# Barriers and Facilitators to Integrating Traditional Chinese Medicine into Primary Healthcare in Singapore: A Systematic Review Protocol

**DOI:** 10.1101/2025.10.01.25337044

**Authors:** Ravi Shankar, Fiona Devi, Xu Qian

## Abstract

**Background:** The integration of Traditional Chinese Medicine (TCM) into primary healthcare systems represents a significant opportunity to enhance healthcare delivery in multicultural societies. Singapore’s unique position as a modern healthcare hub with deep cultural roots in traditional medicine provides an important context for examining this integration process.

**Objective:** To systematically identify, evaluate, and synthesize evidence regarding barriers and facilitators affecting TCM integration into Singapore’s primary healthcare system through a comprehensive mixed-methods systematic review.

**Methods:** This systematic review protocol follows PRISMA-P guidelines and employs a mixed-methods approach. Database searches will include PubMed, Scopus, Web of Science, CINAHL, AMED, Singapore Medical Journal archives, and Chinese medicine databases (CNKI, Wanfang Data) from inception to December 2025. No language restrictions will be applied. Study selection and data extraction will be conducted by two independent reviewers using Covidence systematic review software. Quality assessment will utilize the Mixed Methods Appraisal Tool (MMAT) for diverse study designs, with additional tools (CASP checklists, GRADE-CERQual) applied as appropriate.

**Data Synthesis:** A convergent integrated approach will combine narrative synthesis with framework analysis. The Consolidated Framework for Implementation Research (CFIR) will guide the organization and interpretation of barriers and facilitators across five domains: intervention characteristics, outer setting, inner setting, characteristics of individuals, and process. Quantitative findings will be synthesized narratively, while qualitative data will undergo thematic synthesis. Integration will occur through triangulation protocols and matrix mapping.

**Expected Outcomes:** Comprehensive mapping of multi-level barriers and facilitators, identification of evidence gaps, development of a conceptual framework for TCM integration, and evidence-based recommendations for policy and practice in Singapore’s healthcare system.

## Introduction

The integration of traditional medicine systems into modern healthcare represents one of the most significant developments in global health policy over the past two decades [1]. Traditional Chinese Medicine, with its comprehensive theoretical framework and extensive empirical base spanning over two millennia, has garnered particular attention from healthcare systems seeking to expand therapeutic options and address the limitations of conventional biomedicine [2]. Singapore’s healthcare landscape presents a unique context for examining TCM integration, characterized by its advanced medical infrastructure, multicultural population, and historical acceptance of both Western and traditional medical practices [3].

The Singapore healthcare system has undergone substantial transformation since independence, evolving from a basic public health infrastructure to one of the most advanced healthcare systems in Asia [4]. This evolution has included progressive policies toward traditional medicine, with the establishment of the Traditional Chinese Medicine Practitioners Board in 2001 marking a pivotal moment in the formalization and regulation of TCM practice [5]. Despite these regulatory advances, the actual integration of TCM into primary healthcare settings remains complex and multifaceted, influenced by numerous factors ranging from practitioner training and inter-professional collaboration to patient preferences and healthcare financing mechanisms [4].

Primary healthcare serves as the foundation of Singapore’s healthcare system, following the principles of accessibility, comprehensiveness, continuity, and coordination [6]. The potential integration of TCM into this primary care framework offers opportunities for enhanced patient-centered care, particularly for chronic disease management, preventive health, and conditions where conventional treatments have shown limited efficacy [7]. However, the process of integration faces substantial challenges related to epistemological differences between traditional and biomedical paradigms, standardization of practices, evidence requirements, and professional boundary negotiations [8].

Recent demographic shifts in Singapore, including population aging and the rising burden of chronic diseases, have intensified interest in integrative healthcare approaches that can leverage both conventional and traditional medicine strengths [9]. The COVID-19 pandemic has further highlighted the potential contributions of TCM to public health, with several Singapore institutions exploring TCM-based interventions for pandemic response and recovery [10]. These developments underscore the timeliness and importance of systematically examining the factors that facilitate or hinder effective TCM integration into primary healthcare settings.

The conceptualization of integration itself requires careful consideration, as it encompasses various models ranging from simple co-location of services to fully integrated care pathways with shared clinical decision-making [11]. Singapore’s approach has primarily focused on regulated coexistence rather than systematic integration, with TCM practitioners operating largely in parallel to conventional primary care providers [12]. Understanding the barriers and facilitators to deeper integration requires examination of multiple stakeholder perspectives, institutional arrangements, regulatory frameworks, and cultural factors unique to Singapore’s context.

## Background and Rationale

The global landscape of integrative medicine has evolved considerably over the past three decades, with increasing recognition of traditional medicine systems by international health organizations and national governments [13]. The World Health Organization’s Traditional Medicine Strategy emphasized the importance of integrating traditional medicine into national health systems, promoting a vision of universal health coverage that includes access to safe and effective traditional medicine services [1]. This global momentum has influenced healthcare policy discussions in Singapore, where TCM has maintained a significant presence despite rapid modernization and westernization of the healthcare sector [14].

Singapore’s multicultural society, comprising Chinese, Malay, Indian, and other ethnic groups, creates a complex healthcare landscape where different traditional medicine systems coexist alongside Western biomedicine [15]. Among these traditional systems, TCM holds particular prominence due to the Chinese majority population and the systematic development of TCM education and regulation over the past two decades [5]. The establishment of Nanyang Technological University’s double degree program in Biomedical Sciences and Chinese Medicine in 2005 represented a significant step toward bridging traditional and modern medical paradigms through education.

The primary healthcare sector in Singapore operates through a network of polyclinics, family medicine clinics, and community health centers that serve as the first point of contact for most health concerns [16]. These primary care settings handle approximately 45% of all primary care attendances in the public sector and play a crucial role in chronic disease management through programs such as the Community Health Assist Scheme (CHAS) and the Primary Care Networks (PCN) [7]. The potential integration of TCM into these existing structures presents both opportunities and challenges that require systematic investigation.

Traditional Chinese medicine remains widely utilized in Singapore, with national surveys demonstrating substantial engagement with TCM services among the population. The Ministry of Health’s 2010 National Health Survey revealed that a significant proportion of Singaporeans had consulted TCM practitioners, with common reasons including general well-being, musculoskeletal complaints such as sprains, chronic pain conditions including headaches and back pain, and acute minor illnesses [17]. Despite this widespread utilization, communication barriers persist between TCM users and conventional healthcare providers. Research examining patient-physician communication patterns has identified that discussion of TCM use during Western medicine consultations is influenced by multiple factors, with many patients failing to disclose their concurrent TCM use to their general practitioners [18].

The regulatory framework governing TCM in Singapore has evolved to ensure safety and maintain professional standards while preserving traditional practices. The Traditional Chinese Medicine Practitioners Act 2000 established a statutory board to register and regulate TCM practitioners, with registration requirements including recognized qualifications from approved institutions and successful completion of the Singapore TCM Physicians Registration Examination [17]. However, regulatory harmonization between TCM and conventional medicine sectors remains incomplete, particularly regarding collaborative practice arrangements, shared care protocols, and integrated clinical pathways.

Research on TCM integration in Singapore has primarily focused on individual aspects such as practitioner attitudes, patient preferences, or specific clinical applications. Previous studies have identified various barriers including limited communication between patients and physicians regarding TCM use, with factors such as consultation length, comfort levels, and physician knowledge affecting disclosure of concurrent TCM use [18]. Facilitators identified include successful integration models in hospital settings and growing government support through initiatives such as the Traditional Chinese Medicine Research Grant established by the Ministry of Health in 2014 [8].

The economic dimensions of TCM integration merit particular attention within Singapore’s healthcare financing context. While TCM plays a complementary role in the healthcare system, it does not benefit from the same healthcare financing mechanisms as Western medicine [17]. Recent policy initiatives include a 2020 pilot study extending subsidy and MediSave coverage for acupuncture treatments of neck pain and lower back pain in public healthcare institutions, and a January 2025 regulatory sandbox initiative encouraging public healthcare institutions to integrate Western medicine with TCM using acupuncture and Chinese Proprietary Medicines for chronic disease treatment [8], signaling growing recognition of TCM’s potential role in healthcare delivery.

### Theoretical Framework

This systematic review will employ the Consolidated Framework for Implementation Research (CFIR) as the guiding theoretical framework for organizing and interpreting barriers and facilitators to TCM integration [19]. The CFIR provides a comprehensive structure for understanding implementation processes in healthcare settings and has been successfully applied to various integration initiatives globally. The framework’s five major domains offer a systematic approach to categorizing and analyzing the complex factors influencing TCM integration in Singapore’s primary healthcare context.

The first domain, intervention characteristics, encompasses the features of TCM as an intervention being integrated into primary healthcare. This includes the evidence strength and quality supporting TCM practices, the relative advantage of TCM over existing treatments for specific conditions, the adaptability of TCM practices to different primary care settings, the complexity of implementing TCM alongside conventional care, and the costs associated with TCM integration. Within Singapore’s context, this domain particularly addresses how TCM’s holistic philosophy and personalized treatment approaches align or conflict with standardized primary care protocols [20].

The outer setting domain captures the external influences on TCM integration, including patient needs and resources, cosmopolitanism or networking with external organizations, peer pressure from other healthcare systems, and external policies and incentives. For Singapore, this domain is particularly relevant given the government’s Healthcare 2030 vision and regional developments in integrative medicine across Asia [5]. The multicultural nature of Singapore’s population and varying cultural attitudes toward traditional medicine across different ethnic groups represent important outer setting factors that influence integration dynamics [4].

The inner setting domain focuses on the structural, political, and cultural contexts of primary healthcare organizations where TCM integration occurs. This includes structural characteristics such as the physical space and infrastructure needed for TCM services, networks and communications between TCM and conventional practitioners, organizational culture and its receptivity to traditional medicine, implementation climate including organizational support and resources, and readiness for implementation including leadership engagement and available resources. Singapore’s diverse primary care landscape, encompassing public polyclinics, private general practices, and community health centers, presents varied inner settings that may facilitate or hinder integration differently [4].

The characteristics of individuals domain examines how healthcare providers’ and patients’ attributes influence integration success. This includes knowledge and beliefs about TCM among both conventional and TCM practitioners, self-efficacy in providing or coordinating integrated care, individual stage of change regarding acceptance of integration, individual identification with the organization and its integration goals, and other personal attributes such as cultural background and professional training. The domain recognizes that integration success depends significantly on individual actors’ attitudes, skills, and behaviors within the healthcare system [21].

The process domain encompasses the strategies and tactics used to implement TCM integration, including planning activities, engaging appropriate stakeholders, executing the implementation according to plan, and reflecting and evaluating on the integration progress. This domain is particularly important for understanding how different integration approaches have been attempted in Singapore and their relative success or failure. The framework acknowledges that implementation is not a static event but an active process requiring continuous adjustment and refinement based on experience and feedback [22].

### Objectives

This systematic review aims to comprehensively examine the integration of Traditional Chinese Medicine into Singapore’s primary healthcare system through a rigorous synthesis of available evidence on barriers and facilitators operating across multiple system levels. The primary objective encompasses identifying, evaluating, and synthesizing all relevant evidence to provide a nuanced understanding of the complex factors influencing TCM integration, with specific attention to policy frameworks, organizational structures, professional practices, and patient experiences within Singapore’s unique healthcare context. This overarching goal involves examining both successful and unsuccessful integration initiatives to understand what works, for whom, and under what circumstances, thereby providing actionable insights for stakeholders involved in developing and implementing integrated care models. The review will specifically focus on categorizing barriers across macro-level policy and regulatory constraints such as licensing requirements, scope of practice regulations, and reimbursement policies; meso-level organizational challenges including infrastructure limitations, referral system inadequacies, and inter-professional communication gaps; and micro-level obstacles related to individual practitioner competencies, attitudes, and patient preferences and behaviors. Simultaneously, the review will identify and analyze facilitators that have successfully supported or could potentially enhance TCM integration, including enabling policy mechanisms, supportive organizational structures, successful collaborative models, and positive stakeholder experiences, with particular attention to understanding how different facilitators interact synergistically to promote integration and whether certain combinations of supportive factors are particularly effective in the Singapore context. The synthesis will employ the Consolidated Framework for Implementation Research to systematically organize findings and develop a comprehensive conceptual model that illustrates the relationships between different barriers and facilitators, their mechanisms of influence, and their relative importance across different primary care settings in Singapore, ultimately providing evidence-based recommendations for policy development, implementation strategies, and priority areas for future research and intervention.

## Methods

### Study Design

This systematic review will follow a mixed-methods approach to comprehensively capture and synthesize both quantitative and qualitative evidence related to TCM integration in Singapore’s primary healthcare system. The review protocol has been developed in accordance with the Preferred Reporting Items for Systematic Review and Meta-Analysis Protocols (PRISMA-P) guidelines to ensure methodological rigor and transparency [23]. The mixed-methods design is particularly appropriate for this review given the complex, multifaceted nature of healthcare integration, which involves measurable outcomes as well as experiential and contextual factors that are best captured through qualitative inquiry [24].

The review will employ a convergent integrated approach to synthesis, whereby quantitative and qualitative findings will be analyzed separately initially and then integrated at the interpretation stage to provide a comprehensive understanding of the phenomenon [25]. This approach allows for the triangulation of different types of evidence, potentially revealing complementary insights and identifying areas where quantitative and qualitative findings converge or diverge. The integration process will involve transforming quantitative findings into qualitative themes where appropriate, and using qualitative insights to contextualize and explain quantitative patterns [26]. The Consolidated Framework for Implementation Research (CFIR) will serve as the organizing framework for data extraction and synthesis, ensuring systematic coverage of all relevant implementation domains [19].

### Search Strategy

The search strategy has been developed through consultation with a medical librarian and iterative refinement based on preliminary searches to ensure comprehensive coverage of relevant literature. The strategy combines controlled vocabulary terms (MeSH terms and database-specific thesauri) with free-text keywords to maximize sensitivity while maintaining reasonable specificity. The search will be conducted across multiple electronic databases to ensure comprehensive coverage of biomedical, health services, and complementary medicine literature.

The primary databases to be searched include PubMed/MEDLINE for biomedical literature, Scopus for its broad interdisciplinary coverage, Web of Science for citation tracking and emerging research, CINAHL for nursing and allied health perspectives, and AMED (Allied and Complementary Medicine Database) for complementary medicine-specific content. Regional and specialty databases will also be searched, including the Singapore Medical Journal archives, Index Medicus for South-East Asia Region, and Chinese medicine databases such as CNKI (China National Knowledge Infrastructure) and Wanfang Data, recognizing that relevant research may be published in Chinese language journals.

The search terms have been organized into three main concept groups that will be combined using Boolean operators. The first group captures Traditional Chinese Medicine and related concepts including “Traditional Chinese Medicine,” “TCM,” “Chinese medicine,” “acupuncture,” “herbal medicine,” “tuina,” “cupping,” “moxibustion,” and “qigong.” The second group focuses on primary healthcare and related settings using terms such as “primary care,” “primary health care,” “general practice,” “family medicine,” “community health,” “polyclinic,” and “primary care network.” The third group targets integration and related processes through terms including “integration,” “integrated care,” “integrative medicine,” “collaborative care,” “interprofessional,” “complementary medicine,” “barriers,” “facilitators,” “enablers,” and “challenges.” These groups will be combined with Singapore-specific terms to focus the search geographically.

Grey literature will be searched through government websites including the Ministry of Health Singapore, Health Sciences Authority, and Traditional Chinese Medicine Practitioners Board websites. Professional organization websites such as the Singapore Medical Association, College of Family Physicians Singapore, and Singapore Chinese Physicians’ Association will also be searched. Additionally, ProQuest Dissertations and Theses Global will be searched for relevant doctoral and master’s theses, and conference proceedings from relevant professional meetings will be reviewed. The search will include publications from database inception through December 2025 to ensure the most comprehensive and current evidence base.

### Eligibility Criteria

The inclusion criteria have been developed using the PICOS framework (Population, Intervention, Comparison, Outcomes, Study design) adapted for systematic reviews of complex healthcare interventions. Studies will be included if they focus on the Singapore healthcare context or provide directly applicable insights to Singapore’s healthcare system. The population of interest includes all stakeholders involved in or affected by TCM integration into primary healthcare, including healthcare providers (both TCM and conventional), patients, administrators, and policymakers.

The phenomenon of interest encompasses all aspects of TCM integration into primary healthcare settings, including but not limited to collaborative care models, referral systems, shared care protocols, co-location of services, integrated clinical pathways, and professional education initiatives. Studies examining barriers to integration at any level (policy, organizational, professional, or patient) will be included, as will studies identifying facilitators or enablers of integration. Both implemented integration initiatives and proposed or theoretical models will be considered.

The review will include primary research studies of all designs including randomized controlled trials, quasi-experimental studies, observational studies (cohort, case-control, cross-sectional), qualitative studies (interviews, focus groups, ethnographies), mixed-methods studies, and case studies. Secondary research including systematic reviews, scoping reviews, and narrative reviews will be included for bibliography searching but their findings will be reported separately. Policy documents, guidelines, and grey literature that provide substantive analysis or evidence regarding TCM integration will also be included. No language restrictions will be applied to ensure comprehensive coverage of all relevant literature.

Studies will be excluded if they focus solely on TCM efficacy or effectiveness without addressing integration aspects, examine traditional medicine systems other than TCM without relevance to TCM integration, focus on secondary or tertiary care settings without applicability to primary care. Opinion pieces, editorials, and letters without empirical data or substantive analysis will be excluded, though they may be reviewed for contextual understanding.

### Study Selection

The study selection process will follow a two-stage screening approach to ensure systematic and reproducible identification of eligible studies. All identified citations will be uploaded to Covidence systematic review software for screening and management. Covidence’s automated duplicate detection will be supplemented with manual checking to ensure accuracy in removing duplicate citations. Two independent reviewers will conduct all screening stages, with disagreements resolved through discussion or consultation with a third reviewer when consensus cannot be reached.

The first stage involves title and abstract screening against the predefined eligibility criteria. Reviewers will use Covidence’s standardized screening forms which will be customized for this review and piloted on a sample of 50 citations to ensure consistency. The screening form will include explicit criteria for inclusion and exclusion, with reviewers required to document reasons for exclusion. Studies will be included for full-text review if they potentially meet inclusion criteria or if eligibility cannot be determined from the title and abstract alone. This inclusive approach at the initial screening stage minimizes the risk of inappropriately excluding relevant studies.

The second stage involves full-text screening of potentially eligible studies using Covidence’s full-text review functionality. Full texts will be obtained through institutional library access, interlibrary loans, or direct contact with authors when necessary. Two reviewers will independently assess each full text against the detailed eligibility criteria, documenting specific reasons for exclusion using Covidence’s standardized categories. The software will automatically track conflicts between reviewers and facilitate resolution discussions. A PRISMA flow diagram will be automatically generated by Covidence to document the study selection process, including the number of studies identified, screened, assessed for eligibility, and included in the review, along with reasons for exclusion at the full-text stage.

### Data Extraction

A standardized data extraction form will be developed within Covidence and piloted on a sample of five included studies to ensure comprehensive capture of relevant information. The form will be refined based on the pilot testing before full implementation. Two reviewers will independently extract data from all included studies using Covidence’s extraction templates, with the system automatically highlighting discrepancies for resolution through discussion or third-party arbitration. The data extraction will be guided by the CFIR framework domains to ensure systematic capture of implementation-relevant information.

The data extraction form will capture study characteristics including author(s), publication year, study location within Singapore, study design and methodology, sample size and characteristics, and funding sources. For quantitative studies, additional extraction will include outcome measures used, statistical analyses performed, main findings with effect sizes and confidence intervals where available, and subgroup analyses. For qualitative studies, extraction will focus on theoretical frameworks used, data collection methods, analytical approaches, main themes and subthemes identified, and illustrative quotes that capture key concepts.

Specific data related to barriers and facilitators will be extracted systematically according to CFIR domains. For intervention characteristics, data will include evidence quality, relative advantage, adaptability, complexity, and costs. Outer setting data will capture patient needs, cosmopolitanism, peer pressure, and external policies. Inner setting extraction will focus on structural characteristics, networks, culture, implementation climate, and readiness. Individual characteristics data will include knowledge, beliefs, self-efficacy, and stage of change. Process domain data will capture planning, engaging, executing, and reflecting activities. Contextual factors that may influence the transferability of findings will also be extracted, including healthcare setting characteristics, time period of study, and specific policies or programs in place during the study period.

### Quality Assessment

The methodological quality of included studies will be assessed using appropriate tools based on study design, recognizing that different designs require different quality criteria. The Mixed Methods Appraisal Tool (MMAT) version 2018 will be used as the primary quality assessment instrument, as it allows for the appraisal of qualitative, quantitative, and mixed-methods studies using a single tool, facilitating comparison across diverse study designs [24]. The MMAT includes specific criteria for qualitative studies, randomized controlled trials, non-randomized studies, quantitative descriptive studies, and mixed-methods studies.

For studies not adequately covered by the MMAT, additional tools will be employed. The CASP (Critical Appraisal Skills Programme) checklists will be used for specific study types where more detailed assessment is needed [27]. The GRADE-CERQual approach will be applied to assess confidence in qualitative evidence syntheses, considering methodological limitations, coherence, adequacy, and relevance of the data [28]. Policy documents and grey literature will be assessed using the AACODS (Authority, Accuracy, Coverage, Objectivity, Date, Significance) checklist adapted for health policy analysis.

Two reviewers will independently conduct quality assessments within Covidence, with the platform tracking agreement levels and facilitating resolution of disagreements through discussion or third-party review. Quality assessment results will not be used to exclude studies but rather to provide context for interpretation of findings and to identify areas where evidence is particularly strong or weak. A sensitivity analysis will be conducted to examine whether excluding lower-quality studies substantially changes the review findings. The quality assessment findings will be presented in a summary table and narrative synthesis, highlighting patterns in methodological strengths and limitations across the body of evidence.

### Data Synthesis

The data synthesis will employ a convergent integrated approach that combines narrative synthesis with framework analysis to comprehensively integrate findings from diverse study types. The synthesis process will begin with separate analysis of quantitative and qualitative evidence, followed by integration to develop a comprehensive understanding of barriers and facilitators to TCM integration. The CFIR framework will guide the organization of findings, ensuring systematic coverage of all implementation domains and facilitating identification of patterns across studies.

Quantitative findings will be synthesized narratively due to anticipated heterogeneity in study designs, populations, and outcome measures that would preclude meaningful meta-analysis. The narrative synthesis will follow Popay et al.’s (2006) [29] guidance, including developing a preliminary synthesis of findings, exploring relationships within and between studies, and assessing the robustness of the synthesis. Where multiple studies report similar outcomes, summary tables will be created to facilitate comparison, and vote counting based on direction of effect will be used to identify patterns in the evidence. Effect sizes will be reported where available, with particular attention to the magnitude and consistency of effects across studies.

Qualitative findings will be synthesized using thematic synthesis approaches as described by Thomas and Harden (2008) [30]. This will involve line-by-line coding of findings from primary studies, organization of codes into descriptive themes aligned with CFIR domains, and development of analytical themes that go beyond the primary studies to generate new interpretive explanations. The synthesis will pay particular attention to divergent findings and will explore potential explanations for inconsistencies across studies. NVivo or similar qualitative data analysis software will be used to manage the coding and synthesis process, with coding reliability assessed through independent double-coding of a subset of studies.

The integration of quantitative and qualitative syntheses will follow a triangulation protocol whereby findings from different study types will be compared and contrasted to identify areas of convergence, complementarity, and divergence. A matrix approach will be used to map quantitative and qualitative findings against CFIR domains and specific barriers and facilitators, allowing for visual representation of the evidence base. The integrated synthesis will develop a conceptual framework that illustrates the relationships between different barriers and facilitators, their mechanisms of influence on TCM integration, and their relative importance across different primary care settings in Singapore. This framework will incorporate temporal dimensions to show how barriers and facilitators may evolve over the implementation process.

## Discussion

The systematic review of barriers and facilitators to TCM integration in Singapore’s primary healthcare system represents a critical contribution to the evolving field of integrative medicine in Asian healthcare contexts. The significance of this review extends beyond academic inquiry to practical implications for healthcare policy, clinical practice, and patient care in Singapore and potentially other similar healthcare systems. The comprehensive examination of integration challenges and opportunities will provide evidence-based insights for stakeholders working to develop more inclusive and effective healthcare delivery models. The use of the Consolidated Framework for Implementation Research provides a robust theoretical foundation for understanding the multi-level factors influencing integration success, offering a structured approach to identifying intervention points for improving integration outcomes.

The theoretical implications of this review relate to advancing understanding of healthcare integration as a complex adaptive process influenced by multiple interacting factors across different system levels. The application of CFIR to TCM integration represents a novel contribution to implementation science, potentially revealing unique dynamics in integrating traditional medicine systems that differ from typical healthcare interventions. The review will contribute to theoretical frameworks for understanding how traditional and modern medical systems can coexist and potentially synergize within contemporary healthcare structures. The findings may challenge or refine existing integration models developed primarily in Western contexts by illuminating unique dynamics present in Asian healthcare settings where traditional medicine maintains stronger cultural legitimacy and practical relevance.

The practical implications for healthcare policy in Singapore are substantial, as the review will provide policymakers with comprehensive evidence regarding which policy levers are most likely to facilitate successful integration and which regulatory barriers require attention. The identification of successful integration initiatives and their enabling factors will offer models that could be scaled or adapted for broader implementation. Similarly, understanding persistent barriers will help policymakers prioritize reform efforts and allocate resources more effectively to support integration objectives. The CFIR-guided analysis will particularly highlight the importance of outer setting factors such as policy incentives and regulatory frameworks in shaping integration possibilities.

For healthcare practitioners, both TCM and conventional, the review findings will offer insights into professional challenges and opportunities associated with integrated practice. Understanding the perspectives and experiences of practitioners who have engaged in collaborative care will provide practical guidance for those considering or beginning integration efforts. The review may also identify professional development needs and educational priorities to better prepare practitioners for working in integrated care settings. The characteristics of individuals domain within CFIR will be particularly relevant for understanding how practitioner beliefs, knowledge, and self-efficacy influence integration success.

This systematic review is expected to generate several significant outcomes that will advance understanding of TCM integration in Singapore’s primary healthcare system. The primary outcome will be a comprehensive mapping of barriers and facilitators organized according to CFIR domains, providing stakeholders with an evidence-based foundation for developing targeted integration strategies. This mapping will identify which barriers are most consistently reported across studies and contexts, which facilitators show the strongest evidence of effectiveness, and how different factors interact to influence integration outcomes. The review is expected to reveal important patterns in stakeholder perspectives, potentially identifying areas of alignment where collaborative efforts could be most productive, as well as fundamental disagreements that require focused dialogue and negotiation.

A key expected outcome is the identification of evidence gaps that currently limit understanding of TCM integration in Singapore. These gaps may include absence of studies examining specific integration models, lack of economic evaluations of integrated care approaches, limited longitudinal research on integration outcomes, insufficient attention to patient safety in integrated care settings, or inadequate exploration of certain stakeholder perspectives. Identifying these gaps will provide a research agenda for future studies and help prioritize areas where additional evidence is most urgently needed. The CFIR framework will help identify which implementation domains have received insufficient research attention.

The review is anticipated to generate insights into the contextual factors that influence integration success or failure in Singapore’s specific healthcare environment. These insights will be valuable for understanding how Singapore’s unique characteristics including its regulatory framework, multicultural population, healthcare financing system, and professional culture create specific opportunities and challenges for TCM integration. The findings may also identify transferable lessons for other healthcare systems considering similar integration efforts, particularly in Asian contexts with comparable healthcare structures and cultural backgrounds. The systematic application of CFIR will facilitate comparison with integration initiatives in other settings and health systems.

The methodological considerations of conducting this systematic review highlight several important challenges in synthesizing evidence on complex healthcare interventions. The heterogeneity of study designs, outcome measures, and conceptualizations of integration necessitates flexible synthesis approaches that can accommodate diverse evidence types while maintaining methodological rigor. The inclusion of publications in multiple languages addresses potential language bias but also introduces challenges in ensuring consistent interpretation and synthesis across linguistic contexts. The use of Covidence for study management ensures transparency and reproducibility of the review process, setting a standard for future systematic reviews in this field.

The review’s limitations must be acknowledged to appropriately contextualize its findings and conclusions. The focus on Singapore’s specific context, while providing depth and relevance for local application, may limit the generalizability of findings to other healthcare systems. The rapidly evolving nature of healthcare policy and practice means that the review findings will represent a snapshot of current evidence that will require regular updating to maintain relevance. The search cutoff date of December 2025 ensures currency but may exclude very recent developments in this rapidly evolving field. The complexity of the CFIR framework, while providing comprehensive coverage, may make it challenging to synthesize findings in a parsimonious manner.

The potential impact of this review on future research directions is significant, as it will provide a comprehensive baseline assessment of current knowledge and identify priority areas for future investigation. The evidence gaps identified through the review will help researchers focus their efforts on questions most critical for advancing integration practice. The conceptual framework developed through the synthesis may also provide a theoretical foundation for designing and evaluating future integration interventions. The application of CFIR to TCM integration may inspire similar theory-guided approaches to studying traditional medicine integration in other contexts.

## Data Availability

All data produced in the present study are available upon reasonable request to the authors.

